# Two original observations concerning bacterial infections in COVID-19 patients hospitalized in intensive care units during the first wave of the epidemic in France

**DOI:** 10.1101/2021.01.22.21250287

**Authors:** Camille d’Humières, Juliette Patrier, Brice Lortat-Jacob, Alexy Tran-dinh, Lotfi Chemali, Naouale Maataoui, Emilie Rondinaud, Etienne Ruppé, Charles Burdet, Stéphane Ruckly, Philippe Montravers, Jean-François Timsit, Laurence Armand-Lefevre

**Affiliations:** Université de Paris, INSERM, IAME, F-75006 Paris, France; AP-HP, Hôpital Bichat, Bacteriology, F-75018 Paris, France; AP-HP, Hôpital Bichat, Medical and Infectious Diseases ICU (MI2), F-75018 Paris, France; AP-HP, Hôpital Bichat, Department of anesthesiology and surgical critical care, F-75018 Paris, France; Université de Paris, INSERM U 1148, F-75006 Paris, France; INSERM 1152 – ANR-10-LABX-17; AP-HP, Hôpital Bichat, Département d’Epidémiologie, Biostatistique et Recherche Clinique, F-75018 Paris, France

**Keywords:** COVID-19, bacterial infections

## Abstract

Among 197 COVID-19 patients hospitalized in ICU, 88 (44.7%) experienced at least one bacterial infection, with pneumonia (39.1%) and bloodstream infections (15,7%) being the most frequent. Unusual findings include frequent suspicion of bacterial translocations originating from the digestive tract as well as bacterial persistence in the lungs despite adequate therapy.

## Introduction

Since December 2019, the world has been facing a pandemic due to the spread of SARS-CoV-2 coronavirus, which may lead to acute respiratory failure. In 6% of cases, patients are hospitalized[1], and 20% of whom develop acute respiratory distress syndrome for which they are admitted to intensive care unit (ICU)[1]. Hospitalization in ICU is highly associated with health-care infections, particularly ventilator-associated pneumonia (VAP) and bloodstream infections (BSI). In France, the first COVID-19 patients, all from Wuhan (China), were hospitalized at the end of January 2020[2]. One of them, admitted to the medical ICU of our hospital, was surprisingly co-infected upon arrival with an unexpected antibiotic susceptible *Acinetobacter baumannii*. Two meta-analyses reported bacterial infection rates in COVID-19 ICU patients of 8.1%[3] and 14%[4]. But data on bacterial infections in COVID-19 patients are still sparse and not always consistent. Moreover, several studies highlight the over-prescription of antibiotics in COVID-19 patients and the risk of a global increase of antimicrobial resistance[5,6]. It is thus essential to know the incidence and epidemiology of bacterial infections in such patients, in order to manage antibiotic prescriptions.

Our study describes bacterial infections in COVID-19 patients hospitalized in two ICUs of a French referral center hospital.

## Material and methods

This retrospective study was conducted on COVID-19 patients hospitalized in ICUs of Bichat Claude Bernard University Hospital (Paris, France) between January 29^th^ (first patient admission) and May 31^st^ 2020. Demographic data, comorbidities and microbiological data have been retrospectively collected.

### Definitions

A COVID-19 confirmed case was defined by a positive result for SARS-CoV-2 virus, based on a reverse transcriptase-polymerase-chain-reaction (RT-PCR) test on a nasopharyngeal swab or respiratory specimen and/or typical parenchyma infiltrates on a chest CT scan.

Bacterial pneumonia was diagnosed according to clinical IDSA guidelines[7] and the presence of bacteria in a respiratory sample (broncho-alveolar lavage (BAL), plugged telescoping catheter (PTC), tracheal aspiration or sputum).

A BSI was defined by the presence of bacteria in blood cultures (BACTEC-FX, Becton-Dickinson) that was not considered contaminant and led to antibiotic initiation or modification.

Early- and late-onset VAP or BSI were established using a breakpoint of 5 days of mechanical ventilation for VAP and of ICU stay for BSI[8].

### Ethics

The Committee for Research Ethics in Anesthesia and Critical care (CERAR) authorized the study (IRB 00010254-2020-168).

### Statistical analysis

For univariate analysis, categorical data were analyzed using Pearson’s chi-squared analysis or Fisher test, and continuous data were analyzed using non-parametric Wilcoxon test.

All statistical analyses were 2-tailed with a significance level of 5%, and were performed with scipy.stat package from Python.

## Results

Overall, 197 COVID-19 patients were admitted to ICU. Median age was 59 years (IQR 50-68) and sex ratio (M/F) was 3. At admission the median SAPS II was 34 (IQR 25-50). Among patients, 116 (58.9%) had at least one co-morbidity and 81 (41.1%) were overweight (body mass index (BMI) ≥ 25 kg/m^2^). In all, 179 patients (91.0 %) received antibiotics (122/179 introduced before ICU and 57/179 during ICU stay) (table S1).

### All ICU infections

Almost half of patients (88/197, 44.7%) had at least one bacterial infection during their ICU stay; 77 (39.1%) had pneumonia, 31 (15.7%) BSI, 4 (2.0%) urinary tract infection and 1 a surgical site infection (table S1). Patients with bacterial infections were more severe at ICU admission than patients without (SAPS II 45 *vs* 30, p<0.001, intubation rate 92.0% *vs* 44.0%, p<0.001, respectively). Patients with bacterial infections had a longer ICU stay (17 days *vs* 5, p<0.001) and a higher mortality rate in ICU (51.1% *vs* 23.9%, p<0.001).

### BSI

Of the 197 COVID-19 patients, 66 (33,5%) had at least one positive blood culture, among which 31 (15,7%) had BSI (table 1). The remaining cases were considered to have contaminated blood cultures, predominantly with coagulase negative staphylococci (95%). Patients with BSI had higher BMIs (30 *vs* 28, p=0.007), were more severe at ICU admission (SAPS II 44 *vs* 33, p=0.007) and during ICU stay (intubation rate 100% *vs* 59%, p<0.001, mortality rate 61.3% *vs* 31.1%, p=0.003) (table 1).

**Table 1:**
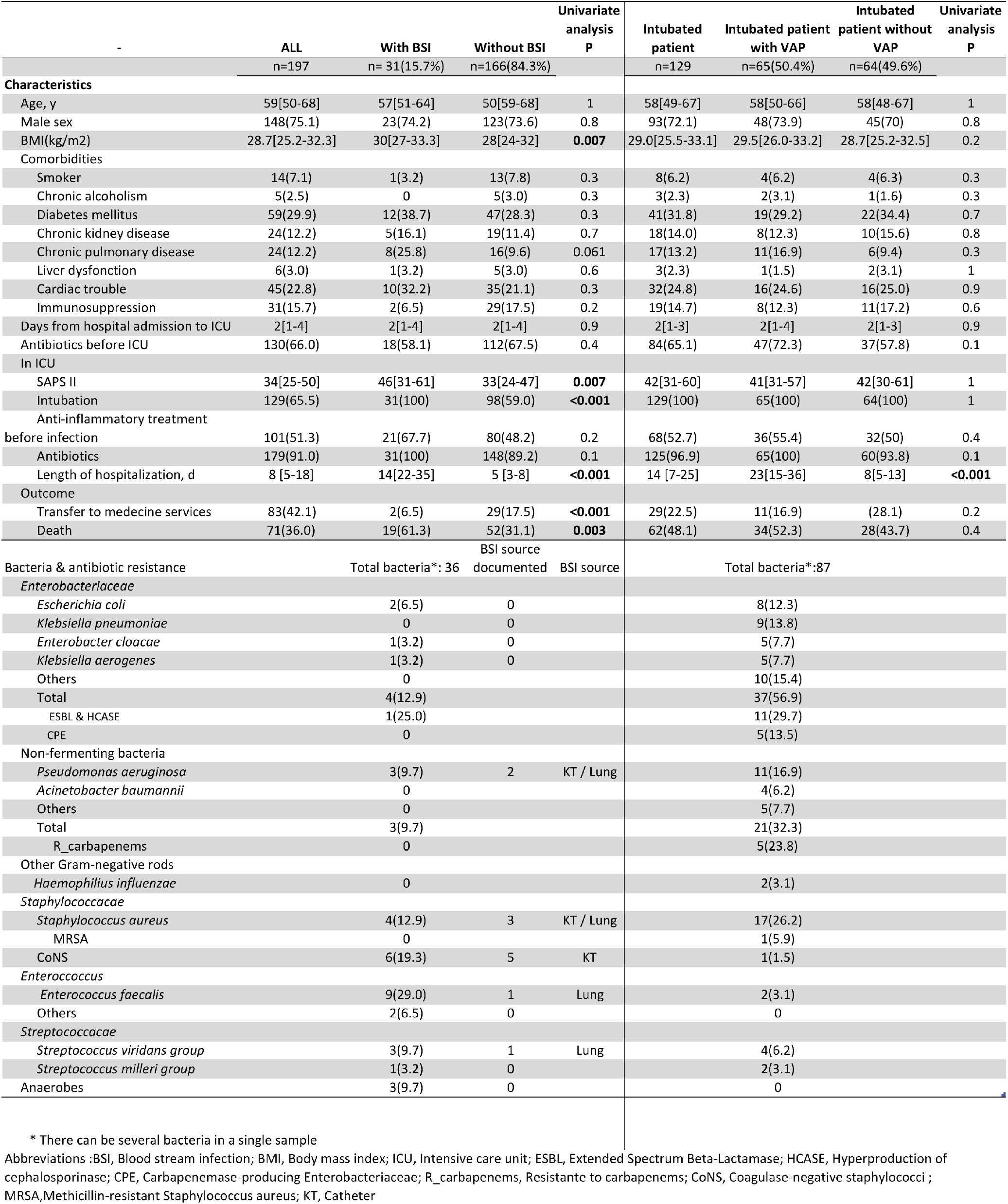
Description of ICU COVID-19 patient with or without bloodstream infections and intubated patients with or without ventilator-associated pneumonia (VAP).

Among the BSI, 83.9% (26/31) were late-onset. Gram-positive cocci were prominent in BSI with 35.5% Enterococci (11/31) and 32.2% Staphylococci (10/31). Enterobacterales were only detected in 4 episodes (4/31, 12.9%) (Table 1).

In 38.7% (12/31) of episodes, the origin of the bacteremia was identified: 5 from the lungs and 7 from catheter (table 1) with positive respiratory sample or catheter tip culture. Of the 19 episodes without proven origin, 17 (17/31, 54.8%) were considered to originate from a digestive or oro-pharyngeal translocation. Indeed, the isolated bacterial species are known to belong to the digestive or oropharyngeal microbiota and all other potential sources were excluded (Table 1).

### Pneumonia

Among the 197 COVID-19 patients, 77 (39.1%) had at least one bacterial pneumonia. Inaugural episodes were: 5 community-acquired pneumonia (6.5%), 7 non-ventilated hospital acquired pneumonia (9.1%) and 65 VAP (84.4%). Median ICU stay and ventilation duration before the first VAP were 10 (IQR, 6-14) and 9 (IQR, 6-14) days respectively; 51/65 (78.5%) VAP were late-onset. The length of hospitalization was longer for patients with VAP than in ventilated patients without VAP (23 *vs* 8 days, p<0.001) (Table 1).

The most frequent bacteria involved in VAP (table 1) were *Staphylococcus aureus* (17/65, 26.2%), *Pseudomonas aeruginosa* (11/65, 16.9%), *Klebsiella pneumoniae* (9/65, 13.8%) and *Escherichia coli* (8/65, 12.3%). No difference was observed in the distribution of bacteria between early- and late-onset VAP (data not shown). Among Enterobacterales, 11/37 (29.7%) were resistant to third-generation cephalosporins but susceptible to carbapenems (8 extended-spectrum beta-lactamase and 3 high-level expression of cephalosporinase) and 5/37 (13.5%) were resistant to carbapenems (all harboring an NDM-type carbapenemase).

Strikingly, we observed an unusual persistence of bacteria in the respiratory samples of patients with VAP despite an adequate antibiotic therapy. Among patients still hospitalized 7 days following the initial VAP, the responsible bacteria was still detected in culture in 57.4% (27/47) of cases. Presence of the bacteria, was 23.1% (6/26) among those who were still hospitalized 16 days following initial VAP (figure S1). The comparison of patients with (27/47) or without (20/47) persistent VAP at D7 shows a lower age (52y *vs* 60.6y, p<0.01) and a higher BMI (31.5 vs 27.8, p=0.018) among those with persistent VAP (Table S2).

## Discussion

Here, we showed that almost half of COVID-19 ICU patients developed a bacterial infection. Our study highlights two bacteriological peculiarities in these patients *(i)* an epidemiology of BSI that may suggest digestive or oropharyngeal translocations and *(ii*) a persistence of bacteria in the lungs of patients adequately treated for VAP.

We observed a higher rate (33.5%) of positive blood cultures than reported in the literature (3.8% to 12%)[9,10] and by the same ICUs last year (12.8%, personal data). More than half of these were considered as contaminants that may be due to changes in guidelines and safety equipment of nursing staff due to the COVID-19 crisis. Bacteria causing BSI were mainly Enterococci (35.5%), Staphylococci (32.2%) and Streptococci (12.9%) which is unusual in ICU patients. Indeed, the bacteria causing BSI in the same ICUs last year were rather Enterobacterales (33.3%), then Staphylococci (30.3%) and Enterococci (18%), (personal data). Interestingly, in 54.8% of episodes, no origin of the BSI could be determined and translocation of bacteria from digestive or oropharyngeal microbiota was suspected. One hypothesis is that the hyper-inflammatory status of COVID-19 patients may increase the permeability of the intestinal or oropharyngeal barriers and thus bacterial translocations. The description of the composition of the intestinal and oropharyngeal microbiota would be helpful to understand the origin of these unusual BSIs[11].

We also observed that 50.4% of COVID-19 ICU intubated patients developed a VAP, a rate much higher than observed in the literature (ranging from 0% in USA[12], 7.5%[13] in Spain, and to 30.6% in France)[14]. The distribution of bacteria causing VAP did not show any particularity, however, we observed a high rate of multi-drug resistant bacteria (MDRB). This could be explained by an increased use of antibiotics in COVID-19 patients (91%), associated with an increase of MDRB dissemination in the ICUs. Surprisingly, we observed that VAP bacteria clearance from the lungs was exceptionally slow despite adequate antibiotic therapy. The cause of antimicrobial failures remains speculative. The main reasons advocated by experts are (i) pharmacodynamic alterations due to frequent glomerular hyperfiltration, even though in our study we did not find any difference in renal function between the patients, (ii) pharmacodynamic alterations due to a high BMI, which seems to be the case in this study, (iii) a probable decrease in the antimicrobial lung concentration associated with pulmonary emboli and obstruction of the pulmonary vasculature frequently observed in ICU COVID-19 patients[15], and/or (iv) an impaired immune response. To our knowledge, these observations have not previously been described in the literature.

This retrospective observational study was conducted in a single center, comprising two ICUs and with a small sample size and our observations should be confirmed by a multi-center case-control study and experimental confirmation.

In conclusion, this work is the first of its kind to describe bacterial persistence in the lungs despite adequate therapy, as well as frequent suspicion of bacterial translocations originating from the digestive or oropharyngeal microbiota, in COVID-19 ICU patients.

## Data Availability

NA

---

**Table S1:** Description of patients COVID-19 in ICU and comparison of patients with or without bacterial infections

|  | All COVID-19<br>patients<br>n=197 | bacterial<br>infection<br>n=88 | no bacterial<br>infection<br>n=109 | Univariate<br>analysis<br>P |
| --- | --- | --- | --- | --- |
| Characteristics |  |  |  | - |
| Age, y | 59[50-68] | 58[50-66] | 59[50-68] | 1 |
| Male sex | 148(75.1) | 67(76.1) | 79(72.5) | 0.7 |
| **BMI(kg/m<sup>2</sup>)** | **28.7[25.2-32.3]** | **29[26-33]** | **28[25-31]** | **0.04** |
| Comorbidities |  |  |  |  |
| Smoker | 14(7.1) | 7(7.9) | 7(6.4) | 0.9 |
| Chronic alcoholism | 5(2.5) | 2(2.2) | 3(2.7) | 1 |
| Diabetes mellitus | 59(29.9) | 27(30.7) | 32(29.4) | 1 |
| Chronic kidney disease | 24(12.2) | 10(11.4) | 14(12.8) | 0.9 |
| Chronic pulmonary disease | 24(12.2) | 15(17.0) | 9(8.3) | 0.1 |
| Liver dysfunction | 6(3.0) | 1(1.1) | 5(4.6) | 0.3 |
| Cardiac trouble | 45(22.8) | 22(25) | 23(21.1) | 0.6 |
| Immunosuppression | 31(15.7) | 13(14.8) | 18(16.5) | 0.9 |
| Days from hospital admission to ICU | 2[1-4] | 2[1-4] | 2[1-3] | 0.8 |
| Antibiotics before ICU | 130(66.0) | 59(67.0) | 71(65.1) | 1 |
| In ICU |  |  |  |  |
| **SAPSII score** | **34[25-50]** | **45[31-63]** | **30[22-42]** | **<0.001** |
| **Intubation** | **129(65.5)** | **81(92.0)** | **48(44.0)** | **<0.001** |
| Anti-inflammatory treatment before infection | 101(51.3) | 49(55.7) | 52(47.7) | 0.4 |
| **Antibiotics** | **179(91.0)** | **88(100)** | **91(83.5)** | **<0.001** |
| **Length of hospitalization, d** | **8[5-18]** | **19[12-31]** | **5[3-8]** | **<0.001** |
| Outcome |  |  |  |  |
| **Transfer to medicine services** | **83(42.1)** | **18(20.5)** | **65(59.6)** | **<0.001** |
| **Death** | **71(36.0)** | **45(51.1)** | **26(23.9)** | **<0.001** |
| Infections details |  |  |  |  |
| Pneumonia | - | 77(87.5) | - | - |
| Bacteremia | - | 31(35.2) | - | - |
| UTI | - | 4(4.5) | - | - |
| SSI | - | 1(1.1) | - | - |

Abbreviations : BMI, Body mass index; ICU, Intensive care unit; UTI, Urinary tract infection; SSI, Surgical site infection

**Table S2:** Description of patients and bacteria of patient with or without a persistent ventilator-associated pneumonia (VAP) at day 7 post initial VAP diagnosis

|  | Intubated patient with VAP | Persistent VAP at day 7 | no Persistent VAP at day 7 | Univariate analysis P |
| --- | --- | --- | --- | --- |
|  | n=47 | n= 27(57.4%) | n=20(42.6%) |  |
| **Characteristics** |  |  |  |  |
| Age, y | 55[49.5-62.0] | 52.0 [44.0; 56.5] | 60.5 [55.0; 65.0] | **<0.01** |
| Male sex | 37(79%) | 21 (78%) | 16 (80%) | 1 |
| BMI(kg/m2) | 29.1 [26.7; 33.1] | 31.5 [27.2; 34.2] | 27.8 [23.7; 30.4] | **0.018** |
| **Comorbidities** |  |  |  |  |
| Smoker | 2 (4.3%) | 2 (7.4%) | 0 (0%) | 1 |
| Chronic alcoholism | 1 (2.2%) | 1 (3.7%) | 0 (0%) | 1 |
| Diabetes mellitus | 11 (23%) | 6 (22%) | 5 (25%) | 1 |
| Chronic kidney disease | 5 (11%) | 3 (11%) | 2 (10%) | 1 |
| Chronic pulmonary disease | 7 (15%) | 4 (15%) | 3 (15%) | 1 |
| Liver dysfunction | 1 (2.1%) | 4 (20%) | 0 (0%) | 1 |
| Cardiac trouble | 10 (21%) | 6 (22%) | 4 (20%) | 1 |
| Immunosuppression | 6 (13%) | 3 (11%) | 3 (15%) | 1 |
| Antibiotics before ICU | 35 (74%) | 19 (70%) | 16 (80%) | 0.45 |
| **In ICU** |  |  |  |  |
| SAPS II | 43.0 [30.0; 62.0] | 45.0 [28.0; 64.5] | 42.0 [30.8; 50.5] | 0.93 |
| Intubation | 47(100%) | 27(100%) | 20(100%) | 1 |
| Anti-inflammatory treatment before infection | 26 (59%) | 16 (62%) | 10 (56%) | 0.69 |
| Antibiotics | 47(100%) | 27(100%) | 20(100%) | 1 |
| Length of hospitalization, d | 28.0 [21.5; 41.5] | 32.0 [25.0; 47.5] | 19.0 [15.0; 37.0] | **0.012** |
| Creatinin (μmol/L) at D7 | 113[58.0-194] | 113 [64.5; 182] | 108 [45.2; 222] | 0.67 |
| GFR (mL/mn/1,73m<sup>2</sup>) at D7 | 61.6[31.9-90] | 61.6 [32.1; 90.0] | 62.8 [25.7; 90.0] | 0.89 |
| **Outcome** |  |  |  |  |
| Transfer to medecine services | 9 (19%) | 6 (22%) | 3 (15%) | 1 |
| Death | 21 (45%) | 9 (45%) | 12 (44%) | 0.97 |
| **Bacteria & antibiotic resistance** |  |  |  |  |
|  | Total bacteria*: 60 | Total bacteria*: 37 | Total bacteria*: 23 |  |
| *Enterobacteriaceae* |  |  |  |  |
| *Escherichia coli* | 8 | 6 | 2 |  |
| *Klebsiella pneumoniae* | 5 | 2 | 3 |  |
| *Enterobacter cloacae* | 3 | 3 | 0 |  |
| *Klebsiella aerogenes* | 4 | 3 | 1 |  |
| Others | 8 | 6 | 2 |  |
| Total | 28 | 20(71.4%) | 8(28.6%) |  |
| ESBL & HCASE | 8 | 6(75%) | 2(25%) |  |
| CPE | 4 | 4(100%) | 0 |  |
| *Non-fermenting bacteria* |  |  |  |  |
| *Pseudomonas aeruginosa* | 7 | 4 | 3 |  |
| *Acinetobacter baumannii* | 4 | 4 | 0 |  |
| Others | 3 | 2 | 1 |  |
| Total | 14 | 10(71.4%) | 4(28.6%) |  |
| R_carbapenemase | 2 | 2 | 0 |  |
| *Other Gram-negative rods* |  |  |  |  |
| *Haemophilus influenzae* | 2 | 1 | 1 |  |
| *Staphylococcaceae* |  |  |  |  |
| *Staphylococcus aureus* | 11 | 6(54.5%) | 5(56.5%) |  |
| MRSA | 1 | 1 | 0 |  |
| CoNS | 1 | 0 | 1 |  |
| *Enterococcus* |  |  |  |  |
| *Enterococcus faecalis* | 1 | 0 | 1 |  |
| Others |  |  |  |  |
| *Streptococcaceae* |  |  |  |  |
| *Streptococcus viridans group* | 2 | 0 | 2 |  |
| *Streptococcus milleri group* | 1 | 0 | 1 |  |

\* There can be several bacteria in a single sample
Abbreviations :BSI, Blood stream infection; BMI, Body mass index; ICU, Intensive care unit; ESBL, Extended Spectrum Beta-Lactamase; HCASE, Hyperproduction of cephalosporinase; CPE, Carbapenemase-producing Enterobacteriaceae; R\_carbapenemase, Resistant to carbapenems with carbapenemase; CoNS, Coagulase-negative staphylococci ; MRSA,Methicillin-resistant Staphylococcus aureus; KT, Catheter

**Figure S1:**
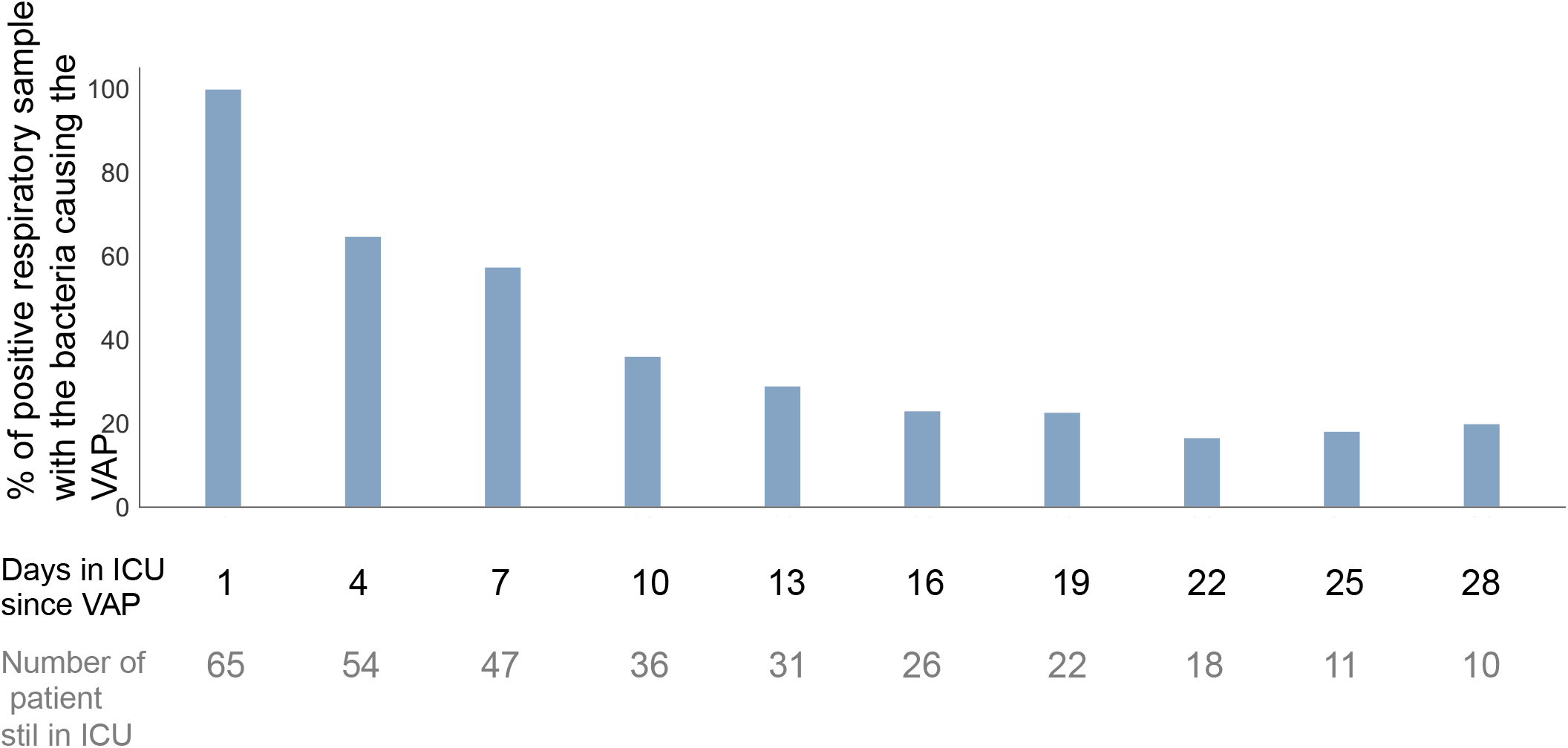
Persistence of bacteria in respiratory samples of COVID-19 patients with VAP

## References

1. Santé Public France. Coronavirus: chiffres clés et évolution de la COVID-19 en France. Available at: https://www.santepubliquefrance.fr/dossiers/coronavirus-covid-19.

2. Bouadma L, Lescure F-X, Lucet J-C, Yazdanpanah Y, Timsit J-F. Severe SARS-CoV-2 infections: practical considerations and management strategy for intensivists. Intensive Care Med 2020; 46:579–582.

3. Langford BJ. Bacterial co-infection and secondary infection in patients with COVID-19: a living rapid review and meta-analysis. Clinical Microbiology and Infection 2020; :8.

4. Lansbury L, Lim B, Baskaran V, Lim WS. Co-infections in people with COVID-19: a systematic review and meta-analysis. Journal of Infection 2020; :S0163445320303236.

5. Rawson TM, Moore LSP, Castro-Sanchez E, et al. COVID-19 and the potential long-term impact on antimicrobial resistance. J Antimicrob Chemother 2020; 75:1681–1684.

6. Huttner BD, Catho G, Pano-Pardo JR, Pulcini C, Schouten J. COVID-19: don’t neglect antimicrobial stewardship principles! Clinical Microbiology and Infection 2020; 26:808–810.

7. Kalil AC, Metersky ML, Klompas M, et al. Management of Adults With Hospital-acquired and Ventilator-associated Pneumonia: 2016 Clinical Practice Guidelines by the Infectious Diseases Society of America and the American Thoracic Society. Clin Infect Dis 2016; 63:e61–e111.

8. Niederman MS. Hospital-acquired pneumonia, health care-associated pneumonia, ventilator-associated pneumonia, and ventilator-associated tracheobronchitis: definitions and challenges in trial design. Clin Infect Dis 2010; 51 Suppl 1:S12–17.

9. Goyal P, Choi JJ, Pinheiro LC, et al. Clinical Characteristics of Covid-19 in New York City. N Engl J Med 2020; 382:2372–2374.

10. Sepulveda J, Westblade LF, Whittier S, et al. Bacteremia and Blood Culture Utilization During COVID-19 Surge in New York City. J Clin Microbiol 2020; :JCM.00875-20, jcm;JCM.00875-20v1.

11. Fontaine C, Armand-Lefèvre L, Magnan M, Nazimoudine A, Timsit J-F, Ruppé E. Relationship between the composition of the intestinal microbiota and the tracheal and intestinal colonization by opportunistic pathogens in intensive care patients. PLoS ONE 2020; 15:e0237260.

12. Bhatraju PK, Ghassemieh BJ, Nichols M, et al. Covid-19 in Critically Ill Patients in the Seattle Region — Case Series. N Engl J Med 2020; 382:2012–2022.

13. Garcia-Vidal C. Incidence of co-infections and superinfections in hospitalised patients with COVID-19: a retrospective cohort study. :25.

14. Dudoignon E, Caméléna F, Deniau B, et al. Bacterial Pneumonia in COVID-19 Critically Ill Patients: A Case Series. Clinical Infectious Diseases 2020; :ciaa762.

15. Ackermann M, Verleden SE, Kuehnel M, et al. Pulmonary Vascular Endothelialitis, Thrombosis, and Angiogenesis in Covid-19. N Engl J Med 2020; 383:120–128.

